# Gender differences in Pornography Use and Associated Factors among high school students: A Cross-Sectional Study

**DOI:** 10.1101/2023.02.07.23285491

**Authors:** Ali Jotyar Mahmoud, Hajar Hassan Abdulqadir, Rojeen Chalabi Khalid, Vindad Hashim Dirbas, Doaa Farhad Hasan, Saman Sherwan Mahfodh, Kareen Yarwant Naisan, Iman Mohammed Arif, Iman Ramadhan Yousif, Sana Najeeb Mohammed, Suzan Muhsin Haji, Suzan Taha Mohammed, Alind Ari Hama Ali, Siavash Babakhani

## Abstract

**Background and aim:** The pornography consuming is increasing through the availability of the internet worldwide and the availability of the pornographic sites without any restriction, pornographic sites have many complications in the consumer and lacking survey in our country which make sense to conduct this study. This study aimed to measure the prevalence of pornography watching among high school students and factors associated with pornography viewing.

**Materials and Methods:** This cross-sectional study was conducted at Zakho independent administration, Kurdistan Region, Iraq among 5 different high school students and 2 institutions. This survey was conducted using a paper questionnaire administered to the participants between April and July 2022. Chi-square analysis was performed to identify possible risk factors for viewing porn and results were expressed as a p value

**Results:** Total number of the participants included in this study was 921 with an average age ± standard deviation (SD) of 16.78 years ±1.26. More than half of participants was male (54.83%), viewed pornography alone (49.08%) and about 69.71% disagree on watching porn. There were statistically significant differences (p <0.001) in the attitude and practice of men compared with women throughout all tested variables. About 50.71% of the participants were viewed pornography at least once in a lifetime, among those 65.1% are male and 34.9% of them are female and 92.55% of participants agree to close pornography sites

**Conclusion:** The prevalence of pornography among high school students is high. Young age, and male gender are predictors for higher pornography viewing and should be considered when designing public health intervention in a related context. Longitudinal studies for investigating pornography consumption among different educational levels are needed to assess the causal relationship between pornography consumption and associated factors.

## Introduction

There is no specific definition for pornography but could be defined any material used to increase the sexual aerosol with increasing in the availability of the internet create a situation which is easy accessibility and cheap to the pornography usage among different age and gender because does not require to print it or downloaded [1]. People watch the pornography due to many causes like get information about the sex, increase excitation during sexual relationships, easier sexual stratification and perform relaxation accordingly handling of this issue is complex [2, 3]. Pornography consumption regard as problem because it is associated with many problems like sexual dysfunction especially at early age [4] and also associated with mental health problem through reducing the memory [5].

There is relationship between the sexual violence and pornography usage [6]. One of the surveys have been done among the young people showed that frequent using of pornography associated with more attraction to the different sexual type like sex as group, with friends and the anal sex [7]. Another research study the link between the violent sex and pornography consumption revealed that those consuming repeatedly for the purpose of sexual violence are associated with other risk factors [8]. A qualitative study showed those men have using pornography their life partners have suffered because they need different types of sex and feeling that the sex is regard physically rather than the love and relationship. In cross sectional study conducted in Arabic countries revealed that men consume pornography more than women [9]. Another study conducted in United states reported that two third (66%) of male participants have viewed pornography at least one time in 12 months while more than one third (39%) of female participants viewed pornography at least one time in the year [10]. Another research have been dine in Australia 87% of the participants have viewed pornography, 99.61% of the male participants viewed pornography at least one time in their life while the female participants 81.7% ever viewing pornography[11]. A survey have been done at Bangladesh 85.07% among those who have encountered pornography are male while only 14.93% of them are female[12] A large survey conducted in Australia have reported 84% of male participants view pornography while 54% of the female view it [13]. The rate of rape and violence in the world also increase in those consuming pornography about 30% (10). The rate of sexually transmitted diseases and hepatitis virus in the world including Iraq also increases among different age groups and gender [14–19]

Many studies have been conducted in Kurdistan Iraq about sexually transmitted diseases including infection causing problem in Genital tract[20>–24] but there is no study about pornography. Despite the increase in pornography consumption during the last decade, pornography viewing differs in accordance with gender and age group [25]. Due to the absence of a large-scale study in Kurdistan region, Iraq, therefore the aim of the present study is to investigate pornography use among high school students and measured gender differences and to determine factors associated with pornography viewing.

## Materials and Methods

### Study Design and participants

This cross-sectional study conducted among high school students in Zakho independent administration Kurdistan-Iraq and used a paper questionnaire administered to the participants between April and July 2022. The total number of the participants enrolled in this study was 921 with the mean age ± standard deviation (SD) of 16.78 years ±1.26. The majority of participants were male 505 (54.83%) (Figure 1).

**Figure 1:**
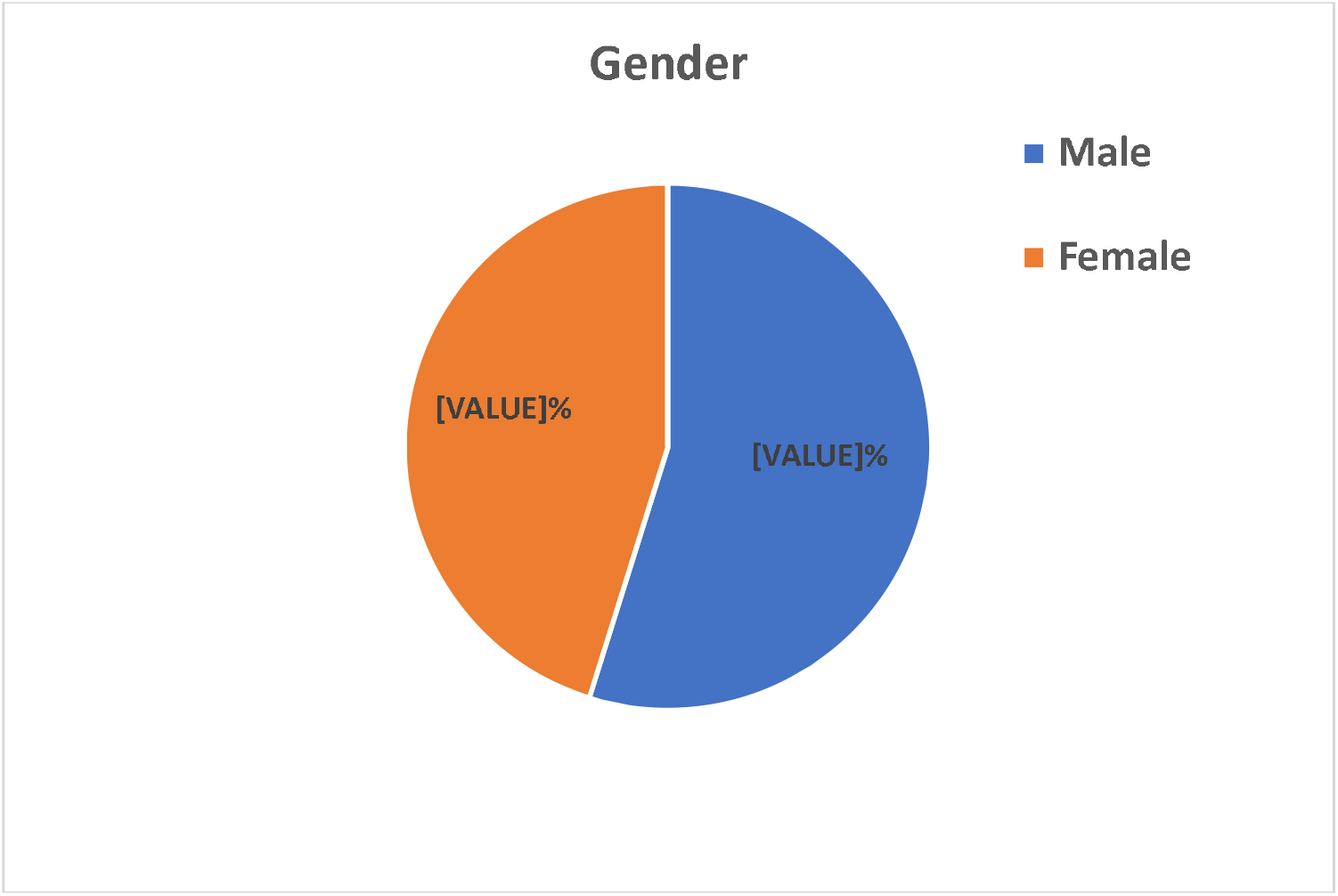
Gender differences use pornography prism.

**Figure 1:**
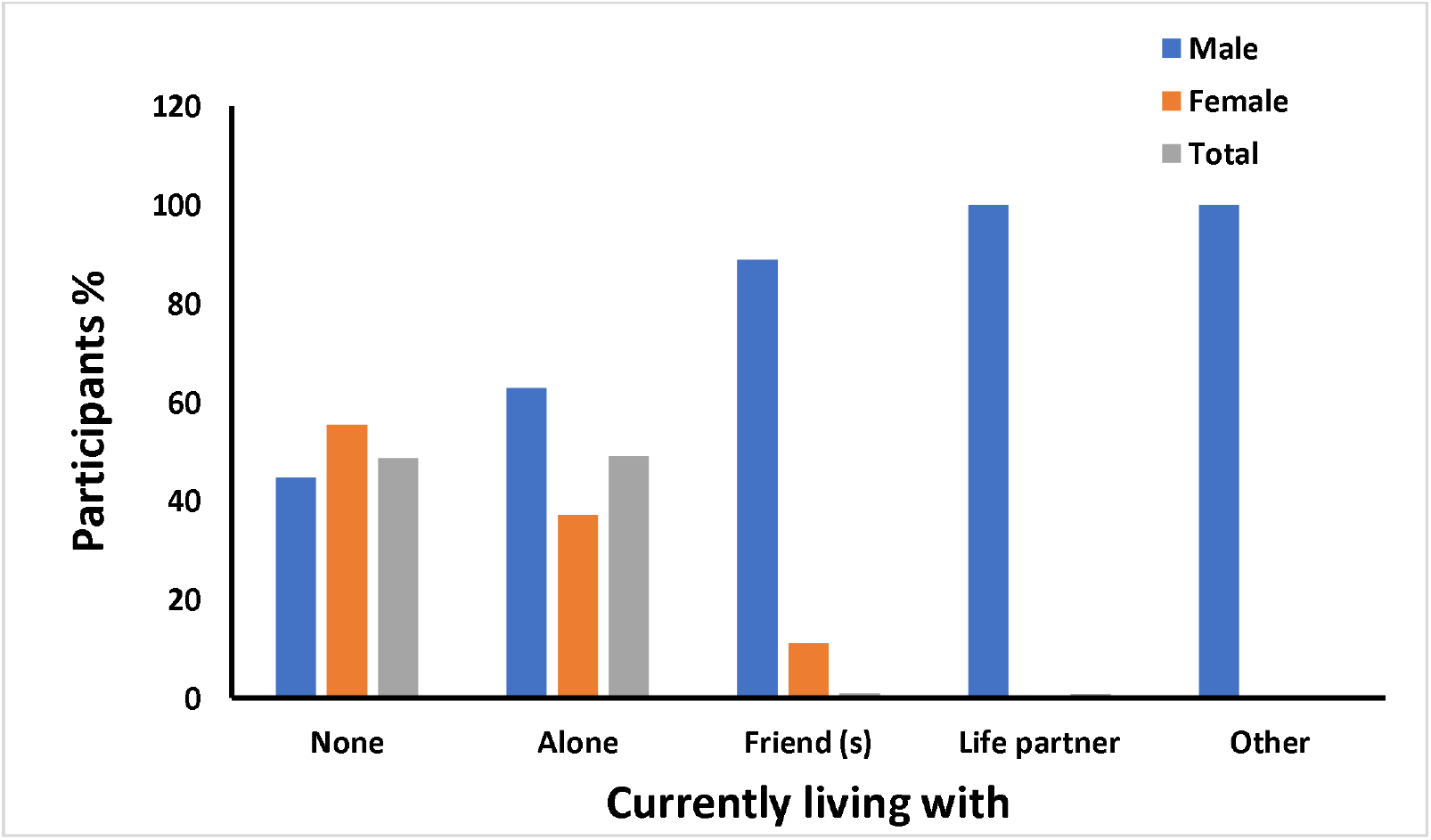
Demographic characteristics of study participants specified by gender.

### Procedure

The questioner has been taken from other survey conducted in Arab countries (1). The questionaries have been printed to be distributed between the students in the different schools because their restriction by the ministry of education that make any smart device forbidden in the schools then we visit each class and filled the questionaries confidentially.

The first part of the questioner about demographic characteristics of participants including gender, address and the age. The second part contained information regarding their first frequency of using, time consumption on pornography and when was the first-time opened pornography and the last part of questionnaire contained information about their idea to the pornography like make pornography forbidden and their agreement about the watching to the pornography. Information from uncompleted survey and the students were not agreed to participate in this study were excluded in the present study

### Ethical consideration

#### Ethical Approval

The protocol, design and procedure of study were approved by the Scientific and Ethics Committee of the College of Medicine, University of Zakho, Duhok Province, Kurdistan Region, Iraq (Ethic committee reference number:4/154/NW). Written informed consent was obtained from all participants before starting analysis

### Statistical analysis

All data were analysed using GraphPad prism version 8.0. Standard descriptive were calculated for each question or item individually. Chi-square test was used to estimate the gender differences with each question individually. P value< 0.05 was considered statistically significant.

## Results

Regarding the agreement of watching pornography, 642 (69.71%) of the participants strongly disagree on watching pornography and more than half of them (51.09%) was female (Table 1). However, 33.44% of participants thought that more than 80% of men view pornography and the majority of them were male (66.23%).

**Table 1.**
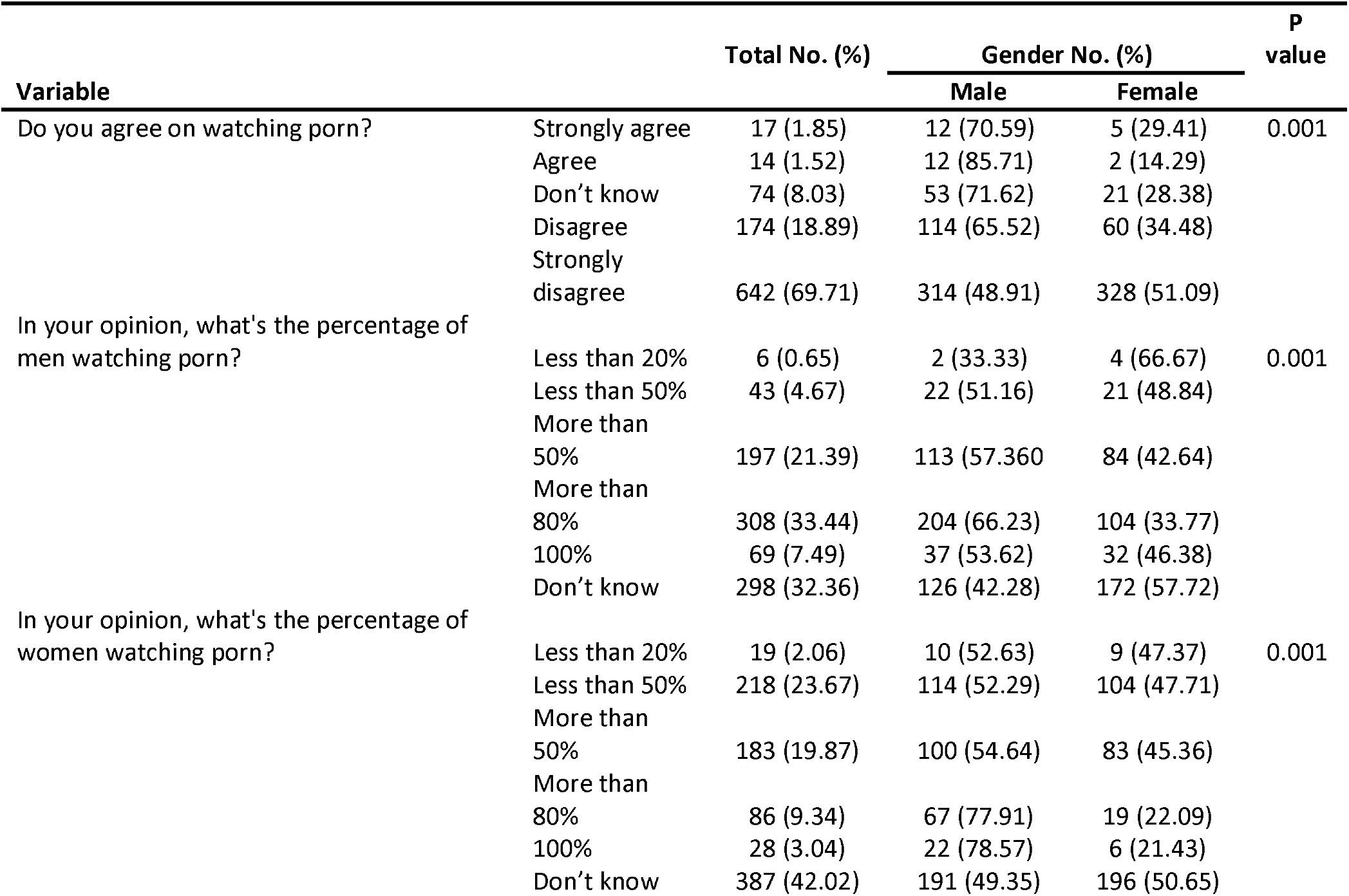
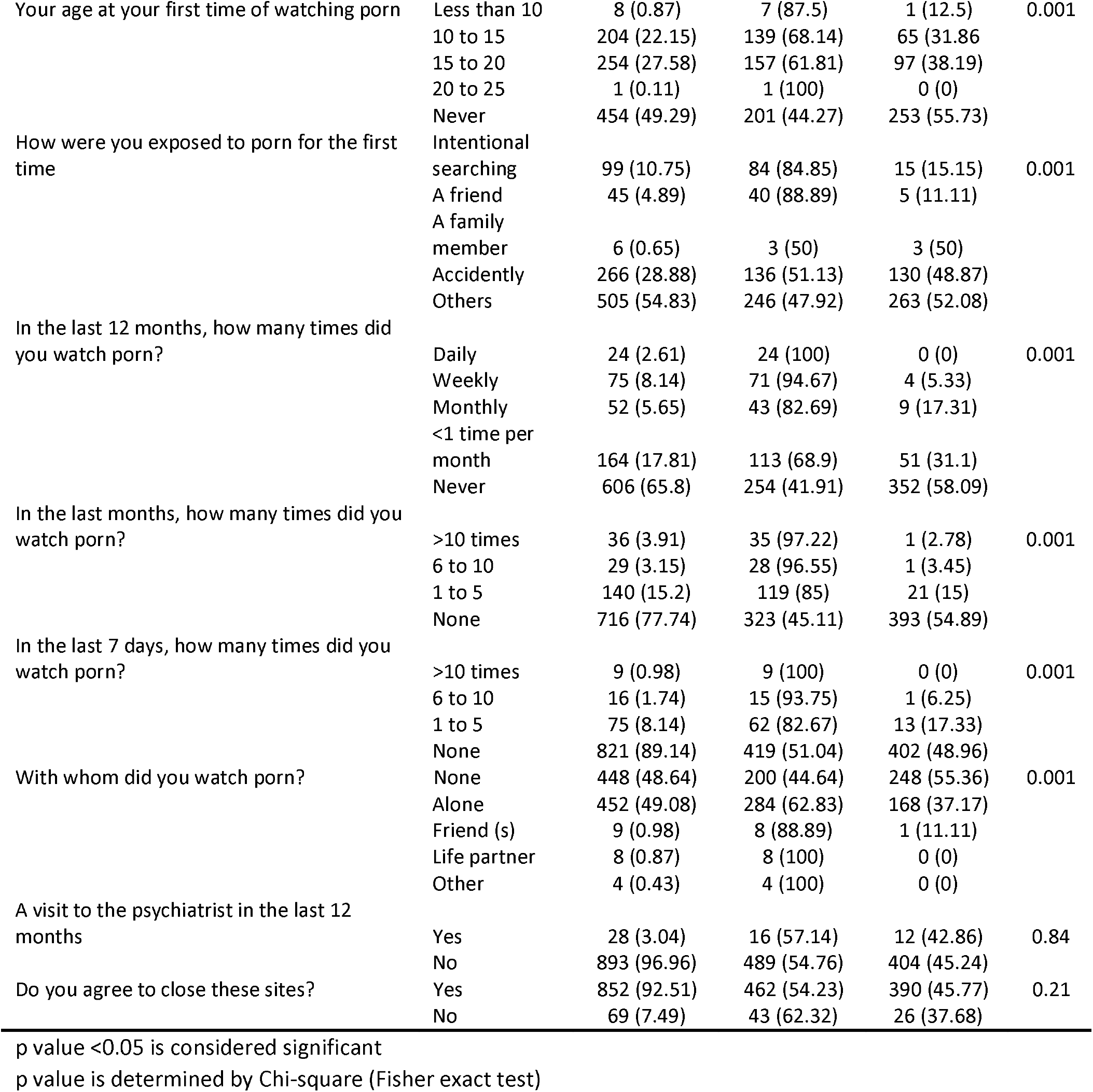
Attitude and practice among gender differences watching pornography.

Regarding their opinion about the percentage of men consumer is 33.44% of participants think that more than 80% of men consume pornography most of them are male, while their opinion about the percentage of female consumer is 40.02% of the participants don’t know, 27.58% of the participants have an experience of consuming pornography between the age of 15-20 years, 28.88% of the participants have exposed to the pornography accidentally and in the 10.75% of the participants have exposed to pornography throw intentional searching from those who see it throw intentional searching (84.85% are male and 15.15 are female), 17.81% of the participants in the last 12 months have consumed pornography less than one time per months, in the last month 15.2% of the participants have consumed pornography between 1-5 time, in the last 7 days 8.14% of the participants consumed it also between 1-5 times, regarding with whom they watched pornography 49.08% of them watch it alone among those who watch it alone (62.83% of them are male and 37.17% of them are female.

## Discussion

According to our knowledge, this the first survey has been conducted in Kurdistan regional Iraq country in order to know how many students in high schools have viewed pornography. Most of the participants were young with a mean age 16.78 of years. Experience of pornography in the last years was 65.8% have never viewed it 17.81% less than one time per month, 5.65% monthly, 8.14% weekly, 2.61% monthly this data is lower than the other survey that have been done like the large multinational in Arabic countries[9] and another one Australia[13], this is because most of our participants belong to the age 15 so may still don’t have an idea about the pornography, our cultural regard the pornography watching shameful and most of the participants don’t believe about anonymously of their ID by this way many of them have bias, most of the participants were Muslim and in Islam watching to the pornography is forbidden. Among those who have consumed pornography 54.39% of them watch it for the first time between the age of 15-20 years, may be this age is higher because of the neurobiological changes in the adolescence age which are increasing in dopamine levels activate penile erection, promote sexual drive with orgasmic quality and increasing of sexual hormone cause increasing in libido [26–28]. It is known that person in this age also have sexual exploration and they want to have sexual experience until they reach to the midtwenties [29, 30]. in our culture sexual activity is forbidden in the outside of the marriage because all the participants are students, they can’t marriage at this age, so they watched pornography that don’t have any restriction increase the chance for viewing it [31, 32]. 69.71% of the participants strongly disagree in watching the pornography this explained that still the community have an awareness their side effects and it is culturally unacceptable. Furthermore, most of the participants have opened these site accidentally which is 57% of all the consumer this is also will increase the chance of addiction, because naturally we have rewarding system in our body for sex and eating for promoting these behavior that is necessary for survival, as the pornography part of the sex the body will have rewarding system for it also [33, 34], so it will increase the numbers of addicted people and the consumers, 54,83% of them choose other this because of the limitation of the questioner the one that have never consumed pornography haven’t another choice according to that the number of the participants that have choose other are 505 and the number of the participants that that haven’t ever viewed these site is 454 by this way the participants that have use another way for watching pornography are only 51. Many of the participants that have consumed pornography are male this also have been reported in other survey[9-13, 35-39], from those who have watched pornography by the intentional searching 84.85% are female and 15.15% are male this is because the female watching the pornography is much more shameful than male watching it. Also, because most of our population are Muslim and some Islamic sects permitting the masturbation for those who don’t have life partners and with low socioeconomic level this make an idea in the population that the semen should be ejaculated frequently00. The present study is cross-sectional design, so the causality can’t be decided by this type of surveys. This subject has not been shared between the students by this way which make the situation hard to understand it and some of them refuse to answer it because they feel it is shameful to talk about pornography. Also, the questioner hasn’t been filled by us because of the confidentiality this also have increase the chance of inaccuracy.

## CONCLUSION

The consumption of the pornography is different according to the age and gender but mostly the man was more consumer than women and the most consumer age are adolescence. This also will increase the chance of infertility and erectile dysfunction. We recommended to be forbidden for those less than 18 years and to have family package for those more than 18 and don’t want to this site being opened even accidentally.

## Data Availability

All data produced in the present work are contained in the manuscript

## Acknowledgment

Authors would like to thank the IFMSA-Kurdistan organisation with their local officer of research exchange Dana S. Abdulkareem, university of Zakho with deanery of medicine collage, directorate of education and security of Zakho independent administration, Kurdistan student’s union, Haval and Zanin institution.

